# Child Mortality in England During the First Year of the COVID-19 Pandemic

**DOI:** 10.1101/2021.08.23.21262114

**Authors:** David Odd, Sylvia Stoianova, Tom Williams, Peter Fleming, Karen Luyt

**Affiliations:** School of Medicine, Division of Population Medicine, Cardiff University, UK; National Child Mortality Database, Bristol Medical School, University of Bristol, St Michael’s Hospital, Southwell Street, Bristol, UK; Centre for Academic Child Health, Population Health Sciences, Bristol Medical School, University of Bristol

**Keywords:** COVID-19, SARS-CoV-2, coronavirus, pandemic, mortality, death, child, infant

## Abstract

**OBJECTIVES:** The aim of this analysis was to quantify the relative risk of childhood deaths across the whole of England during the first year of the COVID pandemic, compared to a similar period of 2019.

**DESIGN:** This work is based on data collected by the National Child Mortality Database (NCMD) which collates data on all children who die in England. The number of deaths, and their characteristics, from 1^st^ April 2020 until 31^st^ of March 2021 (2020-21), were compared to those from the same period of 2019-20. Relative risk and excess mortality were derived for deaths in 2020-21 vs 2019-20.

**SETTING:** All deaths reported to NCMD in England of children under 18 years of age, between April 2019 and March 2021.

**PARTICIPANTS:** 6490 deaths of children, under the age of 18 years, reported to the NCMD over the study period.

**RESULTS:** Children who died between April 2020 and March 2021 had similar demographics to those who died in 2019-20. Overall, there were 356 (198 to 514) fewer deaths in 2020-21 than in 2019-20 (RR 0.90 (0.85-0.94), p<0.001). Repeating the analysis by category of death, suggested that deaths from infection (RR 0.49 (0.38-0.64)) and from other underlying medical conditions (RR 0.75 (0.68-0.82)) were lower in 2020-21 than 2019-20, and weak evidence (p=0.074) that this was also true of deaths from substance abuse.

**CONCLUSIONS:** Childhood mortality in England during the first year of the SARS-CoV-2 pandemic was the lowest on record, with over 300 fewer deaths than the preceding 12 months. The greatest reduction was seen in children less than 10 years old. It is important that we learn from this effect, that potentially offers alternative ways to improve the outcome for the most vulnerable children in our society.

## BACKGROUND

In England, the Office for National Statistics (ONS) published data suggesting an increase in the population mortality of 14% for 2020 compared to previous years^1^. While the direct impact of COVID is significant, the widespread changes in healthcare and society over the year are likely to have had broad impacts on how people accessed healthcare, risks of infectious diseases, and a wider range of social distancing measures. Children have been widely affected by these changes, with the closure of schools, and specific concerns over their mental health^2–4^, the delivery of antenatal care^5,6^, and concerns for vulnerable children; although the direct, infectious, impact of SARS-CoV-2 appears less^7^. Consequently, the overall impact of COVID on childhood mortality is unclear. While some children undoubtably have died as a direct consequence of infection, and others may have suffered due to social restrictions^8–10^, the overall excess mortality for children is unknown. Initial estimates have suggested an overall reduction in child mortality across England^11^, but identifying which children were most affected, whether this affects all demographic groups, and how this relates to the lockdown measures needs clarifying.

This work is based on data collected by the National Child Mortality Database (NCMD) which collates data on all children who die in England before their 18^th^ birthday with death notifications required by statute within 48 hours^12^. The aim of this analysis was to quantify the relative risk of childhood deaths, across the whole of England during the first year of the COVID pandemic, compared to the previous year.

## METHODS

The NCMD commenced data collection on 1^st^ April 2019, and collects data from all 58 Child Death Overview Panels (CDOPs) across England.^13^ All deaths of children in England will ultimately be reviewed in depth by the CDOP panels, but this process often takes many months. In this work, similar to our previous work^11^, to obtain a provisional category of death, all child deaths reported to the NCMD were coded by 3 independent coders (all paediatricians) to identify the most likely category of the cause of death.^14^ All coders recorded a provisional category of death (see below) or that there was insufficient information provided. For each death, if two or more coders agreed on a category this was taken as the most likely category and where no two coders agreed, the category highest in the following hierarchy was used (based on categorisation used by CDOPs).^15^

1. Suicide
2. Substance Abuse
3. Trauma
4. Malignancy
5. Underlying Medical Condition
6. Intrapartum event
7. Preterm Birth
8. Infection
9. SUDIC

In addition, the CDOPs also report baseline characteristics of the child, from which the following data were derived from the notification form:

- Sex of individual (female, male, other (including not known))
- Ethnic Group (Asian or Asian British, Black or Black British, Mixed, Other, Unknown, White)
- Age at death
- The region where the death was reported from
- From the child’s home postcode, the IMD, a measure of local deprivation^16^ (on a score of 1-10) with a lower value suggesting greater deprivation.

From the 1^st^ March 2020, linkage with virology PCR results was performed with Public Health England (PHE) and from April 2020, the Joint Agency Response (JAR) to unexpected child deaths during the COVID-19 Pandemic protocol was amended to include post-mortem viral swabs from all children dying with no immediately identifiable cause.^17^

### Statistical Analysis

Deaths of children occurring from 1^st^ April 2020 until 31^st^ of March 2021 (2020-21) were compared to those from the same period of 2019-20. Initially, we compared the characteristics of those children who died in 2020-21 vs 2019-20. Comparisons were made using Fishers exact test for categorical data, and Mann-Whitney U for age and the IMD deprivation category. Next, using the total number of deaths in the two years the number of excess deaths in 2020-21 was predicted using the 2019-20 data. In addition, the relative risk of deaths between the two years was compared using a Negative Binomial Regression model^4^. Overall population size was assumed to be 12,023,568^18^. This model was then repeated with the deaths split by month of death, and then by demographics of the population: age of death (Less than 1 year, 1-4 years, 5-9 years, 10-14 years or 15-17 years), ethnic group, sex, region of the country, and deprivation score. Models were repeated with the risk calculated for each category, and overall p-values derived to test if the risk of death and year was modified by the characteristic being tested. Population profile was obtained from the 2019 ONS population estimates^18^ for all measures except ethnicity, which was based on the 2011 census data^19^. Absolute numbers of deaths, and excess mortality for 2020-21 vs 2019-20 (with confidence intervals) was also calculated. Finally, we repeated the main model restricting it to each provisional category of death.

Data are presented as median (interquartile range (IQR)), number (%), absolute difference (95% confidence interval (CI)) or relative risk ratio (95% CI). Where frequency counts were below 5, or could be derived, absolute numbers are not presented. Analysis was performed using Stata version 16. All measures had 95% confidence intervals derived. Data was analysed on the 11^th^ of August 2021.

## RESULTS

Data were downloaded from the NCMD system on the 7^th^ of June 2021. Between April 2019 and March 2020 (inclusive) there were 3423 deaths of children, under the age of 18 years, reported to the NCMD. For the corresponding period from April 2020, this had dropped to 3067. Children who died between April 2020 and March 2021 had similar median ages (p=0.215), sex distribution (p=0.939), median deprivation scores (p=0.904) and ethnic groups (p=0.155) to those who died in 2019-20 (Table 1). However, the place of death did appear to be different (p<0.001), with more children dying at home (531 (17.7%) vs 428 (12.7%)) in 2020-21 than 2019-20. A total of 70 children died following a positive SARS-CoV-2 test at any time, with 3 in March 2020, and 67 in the subsequent 12 months.

**Table 1.** Characteristics of all deaths, split by year of death (April to March)

| Measure | Number with Data | 2019 (n=3423) | 2020 (n=3067) | p-value |
| --- | --- | --- | --- | --- |
| Age |  |  |  |  |
| Median (IQR) (Years) | 6490 | 0.2 (0.0-5.3) | 0.1 (0.0-6.2) | 0.215 |
| Sex | 6354 |  |  | 0.939 |
| Female |  | 1466 (43.5%) | 1304 (43.7%) |  |
| Male |  | 1901 (56.5%) | 1683 (56.3%) |  |
| Ethnic Group | 5597 |  |  | 0.155 |
| Asian or Asian British |  | 557 (18.9%) | 462 (17.4%) |  |
| Black or Black British |  | 252 (8.6%) | 247 (9.3%) |  |
| Mixed |  | 188 (6.4%) | 156 (5.9%) |  |
| Other |  | 94 (3.2%) | 65 (2.5%) |  |
| White |  | 1855 (63.0%) | 1721 (64.9%) |  |
| Deprivation Measure (1-10)<sup>16</sup> | 6339 | 4 (2-7) | 4 (2-7) | 0.904 |
| Region | 6490 |  |  | 0.885 |
| East Midlands |  | 283 (8.3%) | 251 (8.2%) |  |
| East of England |  | 339 (9.9%) | 278 (9.1%) |  |
| London |  | 609 (17.8%) | 563 (18.4%) |  |
| North East |  | 154 (4.5%) | 138 (4.5%) |  |
| North West |  | 500 (14.6%) | 431 (14.1%) |  |
| South East |  | 468 (13.7%) | 403 (13.1%) |  |
| South West |  | 252 (7.4%) | 235 (7.7%) |  |
| West Midlands |  | 451 (13.2%) | 436 (14.2%) |  |
| Yorkshire and Humber |  | 367 (10.7%) | 332 (10.8%) |  |
| Place of Death | 6362 |  |  | <0.001 |
| Abroad |  | 10 (0.3%) | 25 (0.8%) |  |
| Home |  | 428 (12.7%) | 531 (17.7%) |  |
| Hospice |  | 145 (4.3%) | 117 (3.9%) |  |
| Hospital |  | 2615 (77.8%) | 2193 (73.1%) |  |
| Other |  | 165 (4.9%) | 133 (4.4%) |  |

Numbers are n (%) or median (IQR) as appropriate
Comparisons were made using Fishers exact or Mann-Whitney U as appropriate

Overall, there were 356 (198 to 514) fewer deaths in 2020-21 than in 2019-20 (RR 0.90 (0.85-0.94), p<0.001) (Table 2). There was evidence that the reduction in deaths was different by age (p<0.001), but no evidence that any other association was significantly modified by month of death (p=0.181), sex (p=0.926), deprivation score (p=0.912), region (p=0.885) or ethnicity (p=0.153). When split by month, fewer deaths appeared to occur in 2020-21 than 2019-20, in October (RR 0.75 (0.63-0.90), November (RR 0.77 (0.65-0.91)) and December (RR 0.80 (0.68-0.94)) (Figure 1). When splitting by age of death, fewer deaths of children below 10 years occurred in 2020-21 than 2019-20 (infants under 1, RR 0.92 (0.87-0.98); 1-4 years, RR 0.67 (0.57-0.78); and 5-9 years, RR 0.81 (0.67-0.98)), but for older children there was no significant reduction in number of deaths (10-14 years, RR 0.95 (0.80 to 1.12); 15-17 years, RR 0.99 (0.85 to 1.15)). When splitting by deprivation, deaths were lower in 2020-21 for all categories except the least deprived (RR 0.93 (0.80 to 1.08)) although there was limited evidence for interaction overall. Similarly when splitting deaths by ethnicity, there was some evidence of a reduction in mortality for all groups except Black or Black British children (RR 0.98 (0.82 to 1.17)), but only limited evidence (p=0.1534) for an overall interaction. There was little change in the relative risks between males and females.

**Table 2.** Relative Risk of Death by calendar month, age, sex, deprivation, region, and ethnicity; with estimates of relative risk between years and the number of excess deaths calculated.

| Measure | Comparison of Deaths between 2019 and 2020 (April to March) |  |  |  |  | *p*<sub>interaction</sub> |
| --- | --- | --- | --- | --- | --- | --- |
|  | 2019 | 2020 | Excess deaths | RR | *p*-value |  |
| All Deaths | 3423 | 3067 | -356 (-514 to -198) | 0.90 (0.85 to 0.94) | <0.001 |  |
| Split by Month of Death |  |  |  |  |  | 0.181 |
| April | 262 | 268 | 6 (-39 to 51) | 1.02 (0.86 to 1.21) | 0.794 |  |
| May | 270 | 269 | -1 (-47 to 45) | 1.00 (0.84 to 1.18) | 0.966 |  |
| June | 238 | 234 | -4 (-47 to 39) | 0.98 (0.82 to 1.18) | 0.854 |  |
| July | 261 | 263 | 2 (-43 to 47) | 1.01 (0.85 to 1.20) | 0.930 |  |
| August | 308 | 263 | -45 (-92 to 2) | 0.85 (0.72 to 1.01) | 0.060 |  |
| September | 275 | 245 | -30 (-75 to 15) | 0.89 (0.75 to 1.06) | 0.189 |  |
| October | 294 | 221 | -73 (-117 to -29) | 0.75 (0.63 to 0.90) | 0.001 |  |
| November | 318 | 245 | -73 (-120 to -26) | 0.77 (0.65 to 0.91) | 0.002 |  |
| December | 333 | 267 | -66 (-114 to -18) | 0.80 (0.68 to 0.94) | 0.007 |  |
| January | 315 | 283 | -32 (-80 to 16) | 0.90 (0.77 to 1.05) | 0.191 |  |
| February | 256 | 237 | -19 (-63 to 25) | 0.93 (0.78 to 1.10) | 0.392 |  |
| March | 293 | 272 | -21 (-68 to 26) | 0.93 (0.79 to 1.09) | 0.377 |  |
| Split By Age |  |  |  |  |  | <0.001 |
| Less than 1 year | 2155 | 1993 | -162 (-288 to -36) | 0.92 (0.87 to 0.98) | 0.012 |  |
| 1-4 years | 394 | 264 | -130 (-180 to -80) | 0.67 (0.57 to 0.78) | <0.001 |  |
| 5-9 years | 243 | 197 | -46 (-87 to -5) | 0.81 (0.67 to 0.98) | 0.029 |  |
| 10-14 years | 289 | 274 | -15 (-62 to 32) | 0.95 (0.80 to 1.12) | 0.527 |  |
| 15-17 years | 342 | 339 | -3 (-54 to 48) | 0.99 (0.85 to 1.15) | 0.908 |  |
| Split by Sex |  |  |  |  |  | 0.926 |
| Male | 1901 | 1683 | -218 (-335 to -101) | 0.89 (0.83 to 0.95) | <0.001 |  |
| Female | 1466 | 1304 | -162 (-265 to -59) | 0.89 (0.83 to 0.96) | 0.002 |  |
| Deprivation Measure* |  |  |  |  |  | 0.912 |
| 1/2 | 1134 | 1012 | -122 (-213 to -31) | 0.89 (0.82 to 0.97) | 0.008 |  |
| 3/4 | 764 | 692 | -72 (-147 to 3) | 0.91 (0.82 to 1.00) | 0.059 |  |
| 5/6 | 616 | 529 | -87 (-153 to -21) | 0.86 (0.76 to 0.96) | 0.010 |  |
| 7/8 | 476 | 413 | -63 (-121 to -5) | 0.87 (0.76 to 0.99) | 0.035 |  |
| 9/10 | 364 | 339 | -25 (-77 to 27) | 0.93 (0.80 to 1.08) | 0.346 |  |
| Region |  |  |  |  |  | 0.885 |
| East Midlands | 283 | 251 | -32 (-77 to 13) | 0.89 (0.75 to 1.05) | 0.166 |  |
| East of England | 339 | 278 | -61 (-110 to -12) | 0.82 (0.70 to 0.96) | 0.014 |  |
| London | 609 | 563 | -46 (-113 to 21) | 0.92 (0.82 to 1.04) | 0.179 |  |
| North East | 154 | 138 | -16 (-49 to 17) | 0.90 (0.71 to 1.13) | 0.349 |  |
| North West | 500 | 431 | -69 (-129 to -9) | 0.86 (0.76 to 0.98) | 0.024 |  |
| South East | 468 | 403 | -65 (-123 to -7) | 0.86 (0.75 to 0.98) | 0.028 |  |
| South West | 252 | 235 | -17 (-60 to 26) | 0.93 (0.78 to 1.11) | 0.441 |  |
| West Midlands | 451 | 436 | -15 (-73 to 43) | 0.97 (0.85 to 1.10) | 0.615 |  |
| Yorkshire and Humber | 367 | 332 | -35 (-87 to 17) | 0.90 (0.78 to 1.05) | 0.186 |  |
| Ethnic Group |  |  |  |  |  | 0.153 |
| Asian or Asian British | 557 | 462 | -95 (-158 to -32) | 0.83 (0.73 to 0.94) | 0.003 |  |
| Black or Black British | 252 | 247 | -5 (-49 to 39) | 0.98 (0.82 to 1.17) | 0.823 |  |
| Mixed | 188 | 156 | -32 (-68 to 4) | 0.83 (0.67 to 1.03) | 0.085 |  |
| Other | 94 | 65 | -29 (-54 to -4) | 0.69 (0.50 to 0.95) | 0.022 |  |
| White | 1855 | 1721 | -134 (-251 to -17) | 0.93 (0.87 to 0.99) | 0.025 |  |

Numbers are frequency counts or relative risk (RR) plus 95% confidence interval (CI) as appropriate
Shaded rows represent periods of National Lockdown
Some demographic data missing, so denominators may vary between analyses
\* Level one is most deprived area, 10 is most deprived

**Figure 1.**
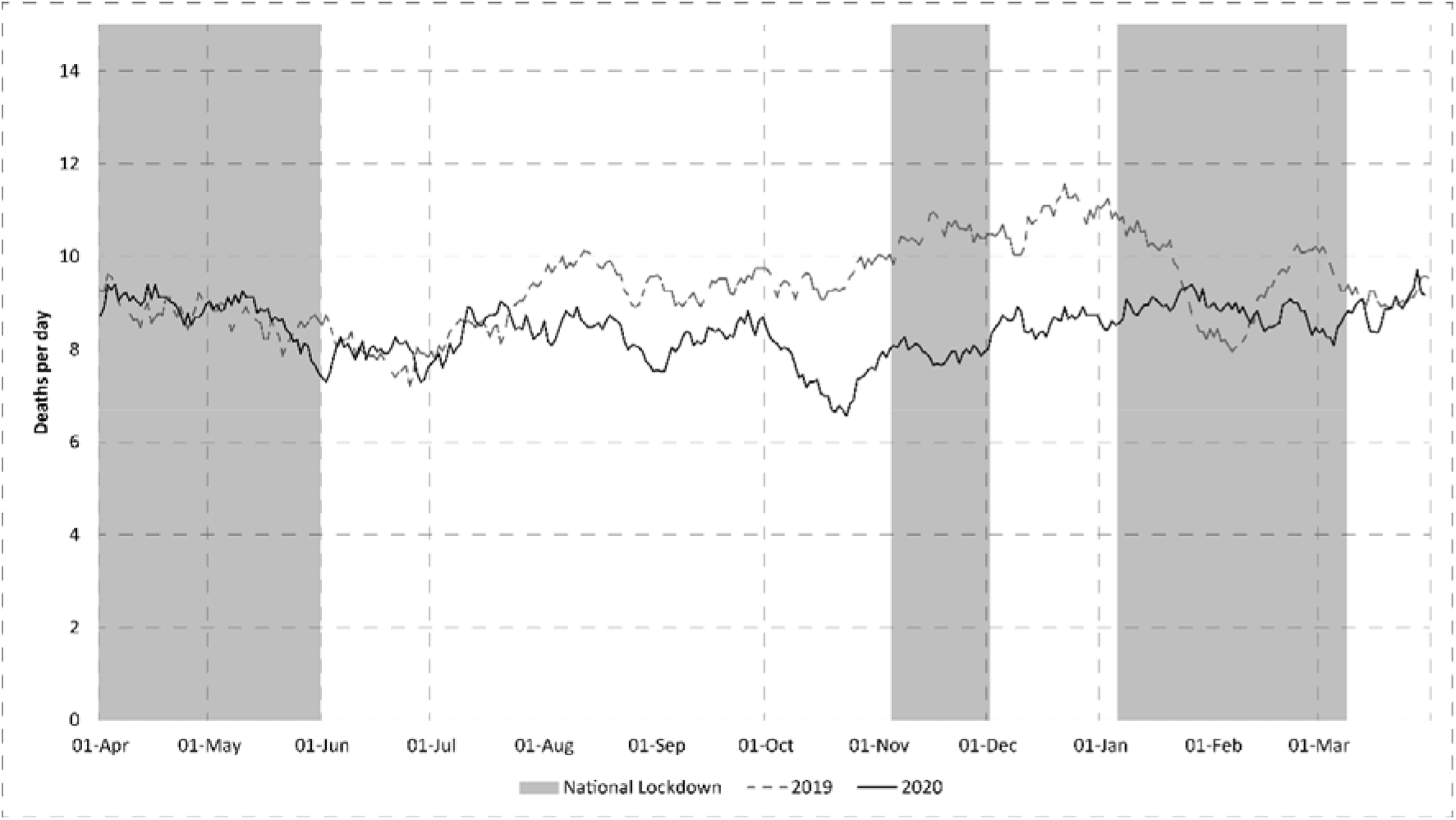
Mean Number of Deaths per day (21 day smoothed (arithmetic) average) split by time period. Periods of National Lockdown Shaded.

Finally, repeating the analysis by category of death, showed that deaths categorised as infection (RR 0.49 (0.38-0.64)) and from other underlying medical conditions (RR 0.75 (0.68-0.82)) were lower in 2020-21 than 2019-20, and there was weak evidence (p=0.086) that this was also true of deaths from substance abuse (Table 3). Mortality from malignancy (p=0.861), preterm birth (p=0.619), intrapartum events (p=0.186), trauma (p=0.102), suicide (p=0.467) and SUDIC (Sudden unexpected deaths in infancy or childhood) (p=0.583) were similar between the two years. Overall, there was strong evidence that the profile of deaths differed between the two years (p<0.001).

**Table 3.** Number of deaths by year (starting April) split by provisional category of death with estimates of relative risk between years and proportion of excess deaths.

| Measure | Comparison of Deaths between 2019 and 2020 (April to March) |  |  |  |  |
| --- | --- | --- | --- | --- | --- |
|  | 2019 | 2020 | Excess deaths | RR | p-value |
| All Deaths | 3423 | 3067 | -356 (-514 to -198) | 0.90 (0.85 to 0.94) | <0.001 |
| Malignancy | 258 | 262 | 4 (-41 to 49) | 1.02 (0.86 to 1.21) | 0.861 |
| Preterm | 902 | 881 | -21 (-104 to 62) | 0.98 (0.89 to 1.07) | 0.619 |
| Intrapartum Events | 166 | 191 | 25 (-12 to 62) | 1.15 (0.93 to 1.42) | 0.186 |
| Infection | 164 | 81 | -83 (-114 to -52) | 0.49 (0.38 to 0.64) | <0.001 |
| Trauma | 164 | 195 | 31 (-6 to 68) | 1.19 (0.97 to 1.46) | 0.102 |
| Substance Misuse | 20 | 10 | -10 (-21 to 1) | 0.50 (0.23 to 1.07) | 0.074 |
| Suicide | 109 | 120 | 11 (-19 to 41) | 1.10 (0.85 to 1.43) | 0.467 |
| SUDIC | 432 | 416 | -16 (-73 to 41) | 0.96 (0.84 to 1.10) | 0.583 |
| Underlying Medical Condition | 1085 | 810 | -275 (-360 to -190) | 0.75 (0.68 to 0.82) | <0.001 |

Numbers are frequency counts or relative risk (RR) plus 95% confidence interval (CI) as appropriate Some deaths had insufficient information to assign a likely category of death

## DISCUSSION

Childhood mortality during the first year of the SARS-CoV-2 pandemic was lower than the preceding year, with over 300 fewer deaths than expected. The reduction in overall mortality was most marked in children less than 10 years of age and appeared due to fewer deaths from infections other than SARS-CoV-2, and fewer deaths of children with other underlying conditions. In addition, the reduction in mortality appeared to occur during the winter months, where the seasonal increase seen in 2019 was not apparent. This period coincided with the prolonged lockdown in England from January to April 2021.

The main limitation of this work is precision of the estimates, due to small numbers. Death in childhood is fortunately a rare event; with absolute mortality across the 2 years of around 27 per 100,000 children per year. While we had adequate precision to identify an overall reduction in deaths, and the likely causal conditions (e.g. infection) in which this occurred, we had less power to identify small, but still potentially important differences that might indicate which children were most affected by the broad social changes, and for which children the effects were less clear. The coincidence of maximum lockdown regulations and the maximum reduction in mortality for younger children suggests that the lockdown regulations including social distancing and reduction of social mixing appeared most beneficial to younger children and perhaps to certain ethnic groups. However, we had limited statistical power and interpretation requires caution. In addition, population measures were derived from ONS data and, particularly in the case of ethnicity, may not be up to date. However, estimates were compared between years and, as the absolute risk is low, error is likely only from population change between the two years. Like all work using routine data, case ascertainment may not be 100%, although reporting to NCMD is a statutory requirement and cohort completeness has previously been reported as good^4^, though we had missing data for some measures (e.g. ethnicity). For this work, the likely cause of death is based on the initial notification data, and the full Child Death Review process may add additional data and detail.^13^

In contrast to emerging data on the mortality seen in adults^20^, the restrictions and adaptations of healthcare delivery placed across England in 2020 to combat COVID resulted in a dramatic reduction of childhood deaths, particularly in the younger children. This effect however is likely to be due to complex processes, and needs to be seen in the context of access to a universal healthcare system, and in low and middle income countries a substantial indirect excess mortality is predicted^21^, alongside substantial broader impacts on the delivery of care^22^, and through impacts on other family members^23^. However, in England, 2020 is likely to be the safest year on record for child deaths; a dramatic reduction from what had been a fairly stable trend over recent years^24^. However, the impact seen here may not be uniform across all groups. While we know that children from some ethnic minority groups appear to be more likely to test positive^11^ and subsequently die^25^ of COVID 19 this pragmatic investigation of both the pandemic, and the responses to it, was unable to confirm a change in the profile of children dying in 2020-21 from the preceding year (other than more deaths occurring at home). However, the reduction in all cause mortality appeared greater for some ethnic minority groups, and in areas of higher deprivation. This is interesting as ethic minority groups^26^ and children in more deprived areas^27^ have recognised higher risk of death than others; and the measures put in place during the first year of the pandemic may have mitigated this patterning. However, it should be noted that for most sub-groups mortality was reduced, and despite the magnitude of the population included, we had small absolute numbers making it difficult to show evidence of a real difference between any sub-groups investigated here, and so results should be interpreted with caution.

There was clear evidence however that the reduction in mortality was seen in two key areas; those children with underlying diseases and those who died of infections. If this reduced mortality in young children was caused by lack of exposure to common and occasionally serious seasonal viral infections such as Influenza, Parainfluenza and RSV, then over the next year or so we may see the effects of increased exposure to these pathogens in vulnerable children at older ages than their usual primary exposure. This may lead to changes in patterns of childhood morbidity from viral infections over the next year. There is, as yet, no clear increase in other causes of death, despite concerns raised around implementation of new ways to deliver healthcare^28^, and concerns over mental health^29^, but impacts may take time to show due to increasing numbers and precision, or through pathways which may take time to impact on mortality (e.g. delayed diagnosis of malignancies or lack of mental health beds or mental health inpatient capacity).

## Conclusion

What these data show is that, during 2020-21, when multiple measures were introduced with the aim of reducing morbidity and mortality from COVID 19 in the adult population, there was an unexpected fall in overall child mortality in England, most marked in younger children and those with underlying health conditions. The magnitude of this fall (around 10%), including those children living in the most deprived conditions, a group for whom previous attempts to reduce excess mortality have generally been less successful, make clear that we need to investigate what aspect of societal reorganisation and the restrictions faced by the whole population have had this effect. It is important that we learn from this effect, that potentially offers alternative ways to improve the outcome for the most vulnerable children in our society.

## Supporting information

Ethics and Consent Statement

Ethics and Consent Statement

## Data Availability

Data is available to approved parties through application to NCMD

## Ethics approval and consent to participate

The National Child Mortality Database (NCMD) is set up as a part of statutory duty to record child mortality, with the purpose to investigate health issues in the child population in order to improve population health. The real-time surveillance of child mortality during the COVID-19 pandemic is delivered by the NCMD programme, with the purpose to investigate an outbreak/incident to help in disease control and prevention. This work is classified as Usual Practice in Public Health and Health Protection (https://eur03.safelinks.protection.outlook.com/?url=http%3A%2F%2Fwww.hra-decisiontools.org.uk%2Fresearch%2Fdocs%2Fdefiningresearchtable_oct2017-1.pdf&data=04%7C01%7COddD%40cardiff.ac.uk%7C4b07c87fa2c94aeee74c08d9661101dd%7Cbdb74b3095684856bdbf06759778fcbc%7C1%7C0%7C637653046409058252%7CUnknown%7CTWFpbGZsb3d8eyJWIjoiMC4wLjAwMDAiLCJQIjoiV2luMzIiLCJBTiI6Ik1haWwiLCJXVCI6Mn0%3D%7C1000&sdata=byPZaHhzCBhUdF2u2sIfvVbPc29keBwnCoXi858IFOk%3D&reserved=0) under the UK Policy Framework for Health and Social Care Research, and was reviewed by the Chair of the Central Bristol NHS Research Ethics Committee with confirmation that NHS ethical approval was not required.

## Availability of data

Aggregate data may be available on request to the corresponding author, and subject to approval by HQIP.

## Transparency

The lead author (the manuscript’s guarantor) affirms that the manuscript is an honest, accurate, and transparent account of the study being reported; that no important aspects of the study have been omitted; and that any discrepancies from the study as planned (and, if relevant, registered) have been explained.

## Authors Contributions

DO: I declare that I participated in the study concept and design, contributed to acquisition, analysis and interpretation of data, drafting and review of the manuscript and that I have seen and approved the final version.

SS: I declare that I participated in the study design, contributed to data acquisition, linkage, analysis and interpretation of analysis, drafting and review of the manuscript; and that I have seen and approved the final version.

TW: I declare that I participated in the study design, contributed to data acquisition, linkage, analysis and interpretation of data analyses, reviewing the manuscript; and that I have seen and approved the final version.

PF: I declare that I participated in the study concept and design, contributed to acquisition and interpretation of data analysis, reviewing the manuscript; and that I have seen and approved the final version.

KL: I declare that I obtained funding for this work, participated in the study concept and design, contributed to data acquisition and interpretation of data, drafting and reviewing the manuscript; and that I have seen and approved the final version.

## Acknowledgements

We thank all Child Death Overview Panels (CDOPs) who submitted data for the purposes of this report and all child death review professionals for submitting data and providing additional information when requested.

Parent and public involvement is at the heart of the NCMD programme. We are indebted to Charlotte Bevan (Sands - Stillbirth and Neonatal Death Charity), Therese McAlorum (Child Bereavement UK) and Jenny Ward (Lullaby Trust), who represent bereaved families on the NCMD programme steering group, for their advice and support with setting up the real-time child mortality surveillance system at the beginning of the COVID-19 pandemic.

Statistical Advice from Professor Chris Metcalfe (University of Bristol).

We thank Dr Yvonne Silove (HQIP) for expert advice around data governance and data sharing for the linkage work.

Public Health England’s Field Service and National Child and Maternal Health Intelligence Network teams, for their collaboration in establishing the real-time surveillance system on child deaths potentially related to COVID-19 and ongoing support in the daily linkage with the SARS_CoV-2 test results.

QES for rapidly developing and deploying the COVID module as part of the NCMD child death notification system.

CleverMed Ltd for their assistance in providing timely neonatal discharge summary data from BadgerNet to support the real-time child mortality surveillance system.

We thank the NCMD team for technical and administrative support.

